# Hilab system, a new point-of-care hematology analyzer supported by the Internet of Things and Artificial Intelligence

**DOI:** 10.1101/2022.01.24.22269469

**Authors:** Aléxia Thamara Gasparin, Claudiane Isabel Franco Araujo, Maiara Carolina Perussolo, Thainá Caroline Schuartz de Jesus, Erika Bergamo Santiago, Ivan Lucas Reis Silva, Ricardo Gurgel de Sousa, Flavia Zhu Teng, Evair Borges Severo, Victor Henrique Alves Ribeiro, Milena Andreuzo Cardoso, Fernanda D’Amico Silva, Carolina Rodrigues de Araujo Perazzoli, João Samuel de Holanda Farias, Bernardo Montesanti Machado de Almeida, Sergio Rogal, Marcus Vinícius Mazega Figueredo

## Abstract

The complete blood count (CBC) is one of the most requested tests by physicians. Mostly realized in conventional hematological analyzers, CBC tests are restricted to centralized laboratories, due to frequent maintenance, size of devices, and expensive costs that these analyzers require. On the other hand, most handheld CBC devices commercially available present high costs and are not liable to calibration or control procedures, which results in poor quality compared to standard hematology instruments. The Hilab system is a small-handed novel hematological platform that uses microscopy and chromatography techniques for blood cells and hematimetric parameters analysis. Combining artificial intelligence, machine learning, and deep learning techniques, provides the main parameters evaluated in the CBC test and four-part differential WBC. For clinical evaluation, accuracy, precision, method comparison, and flagging capabilities of the Hilab System were compared with the Sysmex XE-2100 (Sysmex, Japan) results. Over the entire measuring range, a strong correlation (r > 0.9) between both methodologies was obtained for most parameters evaluated. Also, high accuracy (> 0.85), and adequate precision values were observed. The anticoagulant influence and the sample source (venous and capillary) effect were also evaluated, and no significant differences were observed (p > 0.05). Thus, considering the need for blood count point-of-care tests, especially for quickly patient management, the study indicated that the Hilab system provides fast, accurate, low cost, and robust blood cell analysis for reliable clinical use.

**Key points:**

- The Hilab system presents a high correlation with a standardized reference analyzer for all evaluated CBC parameters;
- The flagging capabilities of the Hilab system present high accuracy compared to results provided by specialized hematologists.

## Introduction

For providing important information about the general patient’s health, the complete blood count (CBC) is one of the most ordered tests by physicians around the world. Besides being used as a screening, it is essential for the diagnosis and evolutionary control of infectious diseases, medical emergencies, surgeries, traumatology, in addition to chemotherapy and radiotherapy accompaniment ^1^ and, therefore, considered an essential diagnostic tool by the World Health Organization ^2^. The main parameters evaluated in CBC are the total count of red blood cells (RBC), platelets (PLT), and white blood cells (WBC). In addition, differential count of white blood cells and the determination of hematimetric parameters, such as hemoglobin (HB), hematocrit (HT), mean corpuscular volume (MCV), and mean corpuscular hemoglobin (MCH), is performed ^3^.

Even considering that the gold standard method for the identification of cells is manual microscopy, currently, CBCs are majority realized in hematological analyzers, which use flow cytometry or resistivity-impedance methodologies ^4^. However, these equipment require frequent maintenance and are large and expensive devices, restricted to hospitals and central laboratories of considerable size. These features decrease the CBC’s access, especially to patients that live far from large urban centers ^5^. In this sense, evaluating the availability of essential diagnostics tools like CBC in different countries of six continents, authors suggest the existence of expressive gaps in diagnostic availability in diverse localities, especially in low and middle-income countries ^6,7^. Besides, the CBC results take a mean time of 24h, which usually promotes a delay in medical diagnosis. Thus, the development of Point-Of-Care technologies (POCT) in hematology allows greater access to exams, in addition to improved medical decisions ^8^.

The Hilab system is a novel hematological platform that uses microscopy and chromatography techniques for blood cells and hematimetric parameters analysis. These small-handed devices are factory calibrated and combine artificial intelligence, machine learning, and deep learning techniques to provide the main parameters evaluated in CBC test and four-part differential WBC. This system is accompanied by single-use test kits that are simple to operate and can be used with venous or capillary samples (containing or not K_3_EDTA). In this study, we evaluated the accuracy of this new POCT, compared to the CBC results provided by Sysmex XE-2100 (Sysmex Corporation, Japan), as well as flagging capabilities, precision studies, and the comparison between venous and capillary blood samples.

## Methods

### 1. Sample preparation process

The Hilab device accepts both venous and fingerprint blood samples. Two drops of blood, totaling 90 uL, are necessary for test realization. The sample is collected directly from the finger (or K3EDTA tube to venous workflow), using the components provided in the test kits. Two single-use diagnostic kits are utilized for the realization of point-of-care CBC tests. The first, used for cell counting, presents a disposable hemocytometer, the diluent solutions, blood collection pipettes, and blood transference pipettes. The second, used for hematimetric parameters evaluation, contains a chromatographic strip and a blood collection pipette (Figure 1; Panel B and E, respectively). Both diagnostic kits present the materials needed for the capillary puncture.

**Figure 1.**
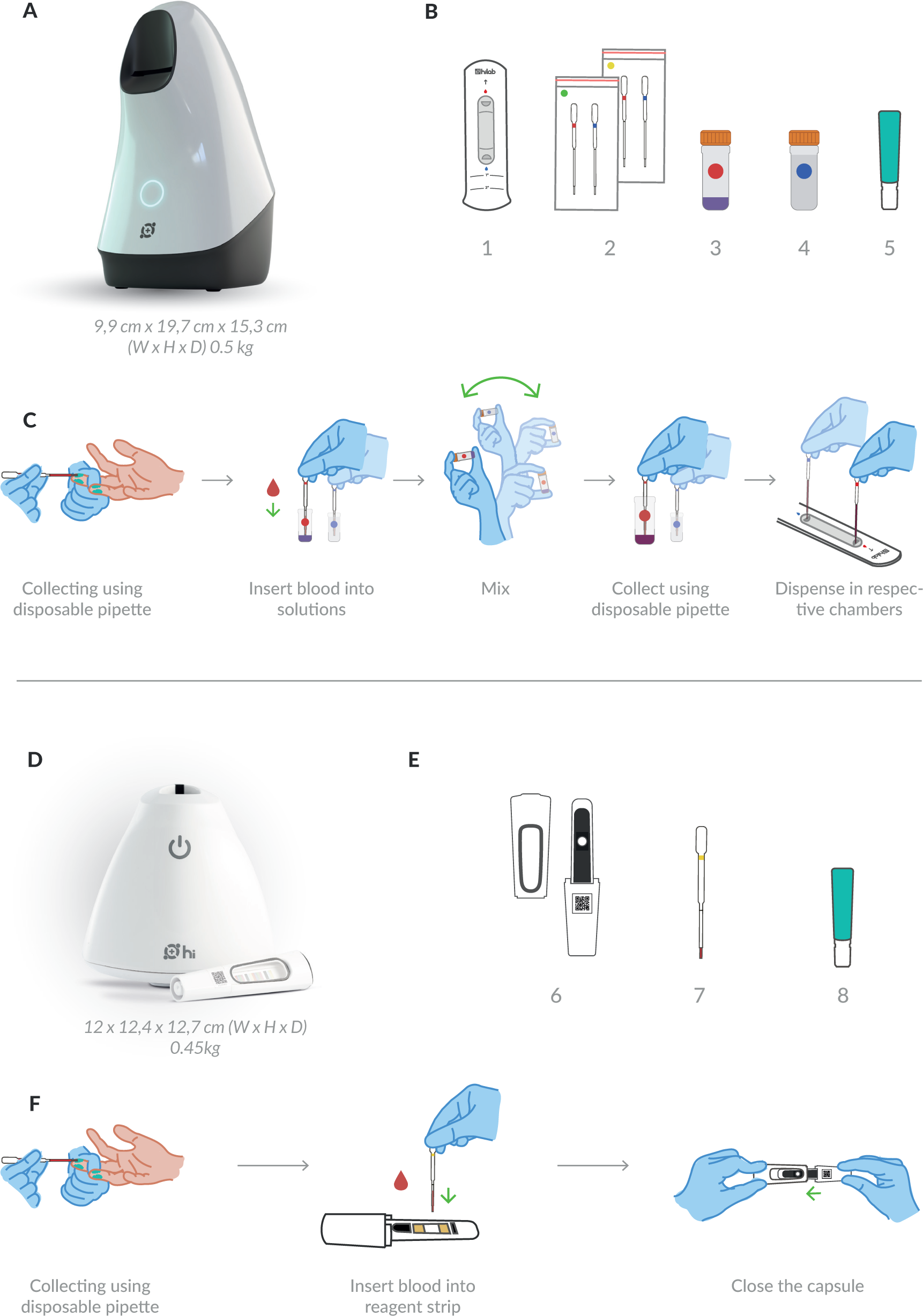
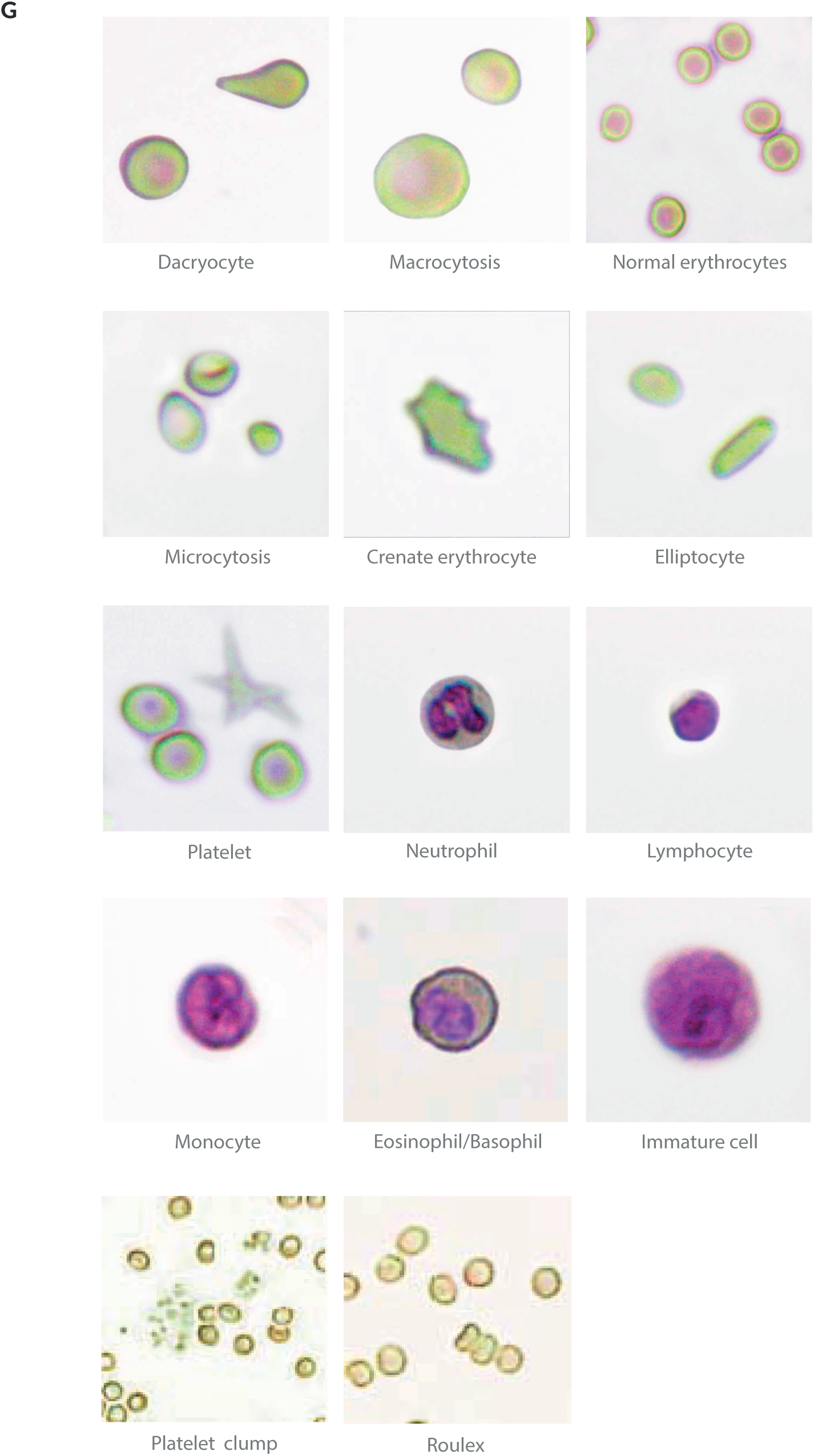
The Hilab system POCT hematology analyzer. Panel A shows Hilab lens device representation (Hilab, Brazil). (B) Components of the Hilab Lens test kit: (1) Hemocytometer, (2) Blood collection pipettes and Blood transfer pipettes, (3) Mixing-bottle of diluent 1; (4) Mixing-bottle of diluent 2; (5) Lancet. (C) Hilab Lens sample preparation workflow for capillary samples. Panel D shows the Hilab Flow device representation (Hilab, Brazil). (E) Components of the Hilab Flow test kit: (1) Capsule; (2) Blood collection pipette; (3) Lancet. (F) Hilab Flow sample preparation workflow for capillary samples. Both single-use test kits accompany isopropyl alcohol swab and curative. (G) Hilab’s software examples of evaluated blood cells.

#### 1.1 Blood Cell count

Two diluent solutions are used to dye the cells and dilute the blood to enable cell counting. During the sample preparation, a drop of blood (40 uL) is collected and placed into solution 1. The same process is done to solution 2. The first diluent composition includes dyes, salts, and surfactants that promote RBC lyse and WBC differential stain. Diluent 2 is composed of different salts, which keep the natural morphology of RBC and PLT. After blood homogenization into solutions, these are individually transferred to the hemocytometer counting chambers (Figure 1; Panel C), allowing the individual blood cell observation in the liquid medium. The process is concluded with hemocytometer insertion into the Hilab Lens device.

#### 1.2 Hematimetric parameters evaluation

For hematimetric parameters evaluation, a single drop of blood (10 uL) is collected and deposited on the chromatographic strip, contained in a plastic capsule. This strip presents a fixed reagent that promotes RBC lysis and HB conversion into methemoglobin. The process is concluded with the insertion of the capsule into the Hilab Flow device for HB quantification (Figure 1; Panel F). From obtained HB values, the artificial intelligence (A.I) estimates the HT, MCV, and MCH, based on previous studies ^9^.

### 2. The device

The Hilab system uses microscopy and chromatography techniques to supply the CBC result. The microscopy one is handled by a small handheld device (19.7 × 9.9 × 15.3 cm; 0.5 kg), called Hilab Lens (Figure 1; Panel A). The acquisition process occurs by the autofocus and image capture process, which takes upwards of 400 images of each sample to form the final image by the composition of all figures stacked. This device processes the blood sample in two stages: the first is used to read the WBC (first chamber), and the second is to read RBC and PLT (second chamber). The chromatography technique is handled by a small handheld analyzer (12.4 × 12.4 × 12.7 cm; 0.45 kg), called Hilab Flow (Figure 1; Panel D). This device incorporates a camera-equipped light detector and samples integrated capsules that enable processing several analytes by the optical density of chromatography strips. Hilab Flow and Lens are calibrated every 24 hours using the calibration capsule, which is used to verify the correct functioning of the sensors and the position of the focus mechanism.

### 3. Imaging acquisition and processing

For Hilab Lens, blood cell images go through a deep learning approach for both cell detection and classification. Data augmentation is applied to RBC, PLT, and WBC subpopulations. This process involves generating more data through rotating and mirroring existing images. The algorithm used for this analysis includes an overlap verification, in which detections that have less than a threshold of intersection are considered the same cell. After detection, images go through feature extraction methods that focus on the shape and texture of the objects to be classified with an independent classifier. For Hilab Flow, HB values are calculated through the regression analysis of colorimetric values. Thus, after the device collects the signals of the blood sample, signal processing techniques extract the mean colorimetric value from the sample. Next, the system applies regression analysis to estimate the HB concentration. Finally, from RBC and HB results, the HCT, MCV, and MCH are estimated.

### 4. Processing of results

After the processing of images by A.I, through Hilab’s software, specialized professionals analyze the exam and issue the report. During the exam processing, if any divergence occurs between the A.I and human results, a senior hematologist also analyzes the exam to achieve the final result. Sample preparation, imaging acquisition, and processing of results by the Hilab system take approximately 30 minutes. In this study, all analyses realized were double-blinded.

### 5. Clinical Protocol

#### 5.1 Method comparison

This study was approved by the Research Ethics Committee of the Paranaense League Against Cancer (CAAE n° 49961421.3.1001.0098). Venous whole blood clinical samples (N = 450) were collected from patients aged between 0.6 and 86 years old, including males (42%) and females (58%), by trained and qualified professionals. The samples encompassed normal and pathological conditions, as thalassemias, anemias, infections, and other blood disorders. The venous whole blood samples were stored in standard K_3_EDTA collection tubes (Vacuette®, Greiner Bio-One, Brazil) and processed within 12h of collection. The sample processing included blood analysis in the Hilab system and the standardized Sysmex XE-2100 analyzer (Sysmex Corporation, Japan), where the reference values were obtained. Pearson correlation, Student-T test, bias, and the Bland-Altman plot of each blood count analyte were calculated and expressed. Also, the Hilab system accuracy, specificity, sensitivity, kappa coefficient, and balanced accuracy were evaluated (confusion matrix; “1” used to values inside normal range and “0” to outside values). All biological samples collected were single-use for this study and discarded after the analysis, following the standard procedure for potentially infected samples. All patients gave written informed consent to participate in this study.

#### 5.2 Precision study

Precision studies were performed using K_3_EDTA whole blood venous samples due to the incompatibility of commercial hematological controls with the Hilab solutions. To encompass the different clinical ranges of CBC analytes, four extended ranges of each PLT, WBC, RBC, and HB sample, were measured ten consecutive times in three distinct devices. For the repeatability study, within-day precision was evaluated by the standard deviation (SD) and the within coefficient of variation (CV) of each range. For the reproducibility study, this protocol was performed by two different operators and evaluated for three consecutive days. Similarly, the SD and CV evaluations of each clinical range were done.

#### 5.3 Equivalence between capillary and venous samples

Fresh fingerprint blood samples were collected from healthy volunteers (n = 150) parallelly with venous blood collection. For all cell blood counts, results from capillary samples were compared with the respective venous plus anticoagulant (K_3_EDTA) samples using the Passing-Bablok analysis and paired Student T-test.

#### 5.4 Flagging Study

The flagging capabilities of the Hilab system were compared to the manual microscopy technique, sending the blood smears to the analysis of trained personnel from a support laboratory (Diagnostico do Brasil®, Parana, Brazil). The evaluation of RBC morphological abnormalities, including microcytosis, anisocytosis, and macrocytosis, was realized. The PLT and WBC abnormalities were also evaluated, assessing the presence of platelet clumps and immature cells, respectively. The accuracy, specificity, sensibility, kappa coefficient, and balanced accuracy of each parameter were calculated and expressed.

#### 5.5 Statistical analysis

All data were analyzed and plotted using the R software statistics package analysis. The Kolmogorov-Smirnov normality test was applied to ensure that the data met the criteria for performing the parametric tests. CV, SD, Bias, Student T-test, Bland-Altman, and Passing-Bablok analysis were calculated using this package. The significance level was set at p ≤ 0.05.

### 6. Data sharing statement

For original data and analysis code, please contact.

## Results

### 1. Method Comparison

The comparability study of CBC analytes is shown in Figure 2. A wide range of values were evaluated for RBC (1.89 - 6.23 × 10^6^/mm^3^), HB (8.5 - 18.9 g/dL), HT (25.3 - 54.5%), MCV (61.8 - 96.1 fL), MCH (17.6 - 33.4 pg), PLT (91.0 - 571.0 × 10^3^/mm^3^), WBC (2.6 - 131.8 × 10^3^/mm^3^), neutrophils (NEU; 1.18 - 58.4 ×10^3^/mm^3^), monocytes (MON; 0.12 - 1.4×10^3^/mm^3^), lymphocytes (LINF; 0.77 - 8.7 × 10^3^/mm^3^), and eosinophils/basophils (EOS/BAS; 0 - 16.5 × 10^3^/mm^3^). As result, a high correlation (r ≥ 0.8) between the Hilab system and the Sysmex XE-2100 analyzer was observed for all analytes (r values; RBC - 0.91; HB - 0.95; HT - 0.96; MCV - 0.95; MCH - 0.99; PLT - 0.95; WBC - 0.99; NEU - 0.99; LIN - 0.95; MON - 0.91; EOS/BAS - 0.8). Excepting EOS/BAS count (p < 0.05), for all analytes evaluated the results provided by methodologies were not statistically different (p < 0.05) each other.

**Figure 2.**
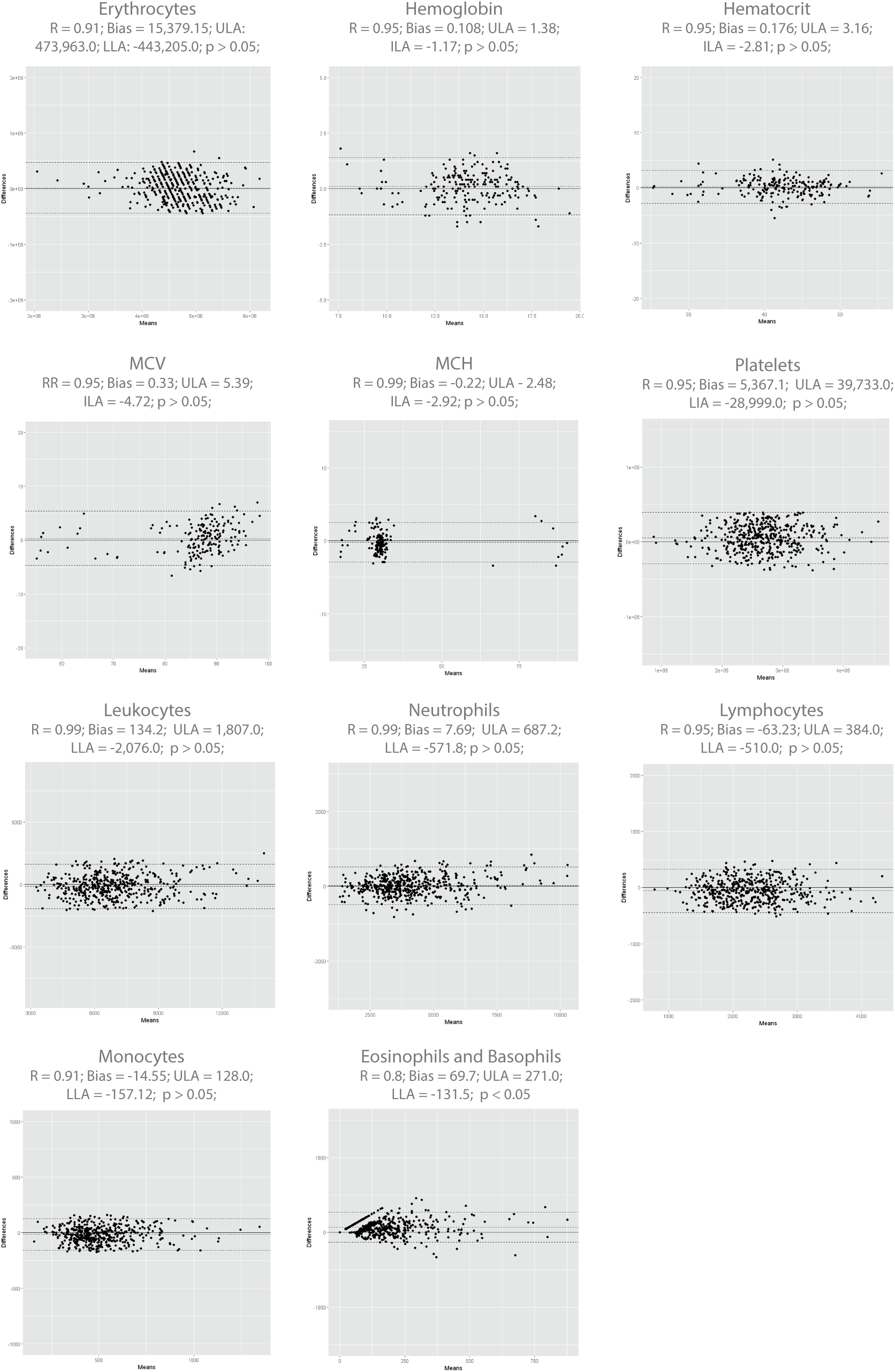
Bland-Altman plot of the method comparison study between the Hilab System and the Sysmex XE-2100. Pearson correlation, bias, Student T-test p-value; upper limit of agreement (ULA), and lower limit of agreement (LLA) are demonstrated for each analyte.

The accuracy, specificity, sensibility, kappa coefficient, and balanced accuracy of each CBC analyte is shown in Table 1. The normal clinical range of RBC (Female (F): 3.9 - 5.1 × 10^6^/mm^3^; Male (M): 4.4 - 5.8 × 10^6^/mm^3^), HB (F: 11.3 - 15.1 g/dl; M: 12.3 - 16.9 g/dl), HT (F: 35.1 - 46.7%; M: 38 - 52.1%), MCV (F: 81 - 100.7 fl; M: 81. 5 - 101.8 fl), MCH (F: 26.3 - 32.6 pg; M: 26.9 - 33.1 pg), PLT (F: 126.6 - 344.7 ×10^3^/mm^3^; M: 128.4 - 302.1 × 10^3^/mm^3^), WBC (F: 2.9 - 10.05 ×10^3^/mm^3^; M: 2.8 - 9.7 × 10^3^/mm^3^), NEU (F: 0.590 - 6.5 × 10^3^/mm^3^; M: 0.550 - 6.35 × 10^3^/mm^3^), MON (F: 0.019 - 0.7 × 10^3^/mm^3^; M: 0,002 - 0.845 × 10^3^/mm^3^), LINF (F: 0.716 - 3.4 × 10^3^/mm^3^; M: 0.582 - 3.4 × 10^3^/mm^3^), and EOS/BAS (F: 0 - 0.574 × 10^3^/mm^3^; M: 0 - 0.718 × 10^3^/mm^3^) were established according previous studies ^10^. As result, all analytes presented high values (≥ 0.8) of accuracy, specificity, sensibility and balanced accuracy. Also, all kappa coefficients were above 0.8 (RBC - 0.94; HB - 0.96; HT - 0.94; MCV - 0.89; MCH - 0.89; PLT - 0.95; WBC - 0.89; NEU - 0.95; LIN - 0.86; MON - 0.95; EOS/BAS - 0.81).

**Table 1.** Accuracy, specificity, sensibility, kappa coefficient, and balanced accuracy of the method comparison study, comparing the Hilab system to Sysmex XE-2100.

|  | **Accuracy (%)** | **Specificity (%)** | **Sensibility (%)** | **Kappa** | **Balanced Accuracy (%)** |
| --- | --- | --- | --- | --- | --- |
| RBC | 99.3 | 93.0 | 99.7 | 0.94 | 96.2 |
| HB | 99.1 | 99.7 | 100.0 | 0.96 | 99.3 |
| HT | 98.7 | 96.3 | 98.9 | 0.94 | 97.6 |
| MCV | 97.6 | 88.5 | 98.9 | 0.89 | 93.7 |
| MCH | 97.0 | 93.6 | 97.6 | 0.89 | 95.6 |
| PLT | 99.8 | 99.9 | 99.8 | 0.95 | 99.0 |
| WBC | 98.0 | 93.5 | 98.6 | 0.89 | 96.0 |
| NEU | 99.4 | 96.5 | 99.6 | 0.95 | 98.07 |
| MON | 99.8 | 99.9 | 99.9 | 0.95 | 99.9 |
| LINF | 99.6 | 99.9 | 99.6 | 0.86 | 99.8 |
| EOS/BAS | 80.0 | 99.0 | 98.0 | 0.81 | 89.1 |

### 2. Precision study

The repeatability and reproducibility studies performed using relevant clinical ranges of PLT, RBC, WBC, and HB, are shown in Table 2. According to European Federation of Clinical Chemistry and Laboratory Medicine guidelines (EFLM; CV variation; PLT < 10%; RBC < 4%; HB < 3.6%; WBC < 15.9%), all parameters presented the CVs inside the established limits (mean CV; RBC - 3.08%; WBC - 6.97%; PLT - 8.16%; HB - 1.63%). The estimated parameters (HT, MCV, and MCH), and the 4-part differential WBC analytes, were not evaluated in these studies.

**Table 2.** Hilab system precision study. Coefficient of variation (CV) and Standard deviation (SD) of each range are demonstrated.

|  | Target range | Repeatability Study |  | Reproducibility Study |  |
| --- | --- | --- | --- | --- | --- |
|  |  | CV (%) | SD | CV (%) | SD |
| **RBC** | 3.0 - 4.0 x 10<sup>6</sup>/mm<sup>3</sup> | 4.15 | 0.152 x 10<sup>6</sup> | 4.10 | 0.148 x 10<sup>6</sup> |
|  | 4.0 - 4.5 x 10<sup>6</sup>/mm<sup>3</sup> | 0.32 | 0.020 x 10<sup>6</sup> | 2.77 | 0.131 x 10<sup>6</sup> |
|  | 4.0 - 5.0 x 10<sup>6</sup>/mm<sup>3</sup> | 3.61 | 0.245 x 10<sup>6</sup> | 2.99 | 0.192 x 10<sup>6</sup> |
|  | 5.0 - 6.0 x 10<sup>6</sup>/mm<sup>3</sup> | 3.04 | 0.156 x 10<sup>6</sup> | 3.70 | 0.390 x 10<sup>6</sup> |
| **WBC** | 1.0 - 2.5 x 10<sup>3</sup>/mm<sup>3</sup> | 10.97 | 0.306 x 10<sup>3</sup> | 9.33 | 0.307 x 10<sup>3</sup> |
|  | 2.5 - 4.5 x 10<sup>3</sup>/mm<sup>3</sup> | 5.64 | 0.412 x 10<sup>3</sup> | 5.83 | 0.200 x 10<sup>3</sup> |
|  | 4.5 - 6.0 x 10<sup>3</sup>/mm<sup>3</sup> | 5.38 | 0.225 x 10<sup>3</sup> | 6.55 | 0.449 x 10<sup>3</sup> |
|  | 6.0 - 7.5 x 10<sup>3</sup>/mm<sup>3</sup> | 6.90 | 0.278 x 10<sup>3</sup> | 5.17 | 0.244 x 10<sup>3</sup> |
| **PLT** | 50 - 90 x 10<sup>3</sup>/mm<sup>3</sup> | 7.24 | 7.0 x 10<sup>3</sup> | 7.70 | 10.5 x 10<sup>3</sup> |
|  | 90 - 150 x 10<sup>3</sup>/mm<sup>3</sup> | 5.78 | 20.5 x 10<sup>3</sup> | 8.93 | 20.3 x 10<sup>3</sup> |
|  | 150 - 250 x 10<sup>3</sup>/mm<sup>3</sup> | 6.50 | 21.9 x 10<sup>3</sup> | 8.80 | 24.3 x 10<sup>3</sup> |
|  | 250 - 400 x 10<sup>3</sup>/mm<sup>3</sup> | 8.66 | 19.15 x 10<sup>3</sup> | 11.70 | 20.2 x 10<sup>3</sup> |
| **HB** | 4.0 - 8.0 g/dL | 1.3 | 0.206 | 2.0 | 0.215 |
|  | 8.1 - 12.9 g/dL | 2.1 | 0.395 | 1.7 | 0.301 |
|  | 13.0 - 15.9 g/dL | 1.6 | 0.289 | 1.0 | 0.190 |
|  | 16.0 - 19.0 g/dL | 1.9 | 0.277 | 1.5 | 0.310 |

### 3. Anticoagulant influence and the effect of sample type

To assess the anticoagulant influence and the effect of sample type for cell analytes, the Hilab system results of whole venous blood were compared with the respective freshly fingerprint blood results (Figure 3). Evaluating the paired Student T test, for all analytes, no statistical differences (p > 0.05) between venous (plus anticoagulant) and capillary blood samples were observed (p values; RBC - 0.90; PLT - 0.39; WBC - 0.76; NEU - 0.51; LIN - 0.92; MON - 0.60; EOS/BAS - 0.67). Besides, all analytes presented high correlation (r > 0.85) between venous and capillary blood samples (r values; RBC - 0.92; PLT - 0.94; WBC - 0.96; NEU - 0.99; LIN - 0.95; MON - 0.96; EOS/BAS - 0.84).

**Figure 3.**
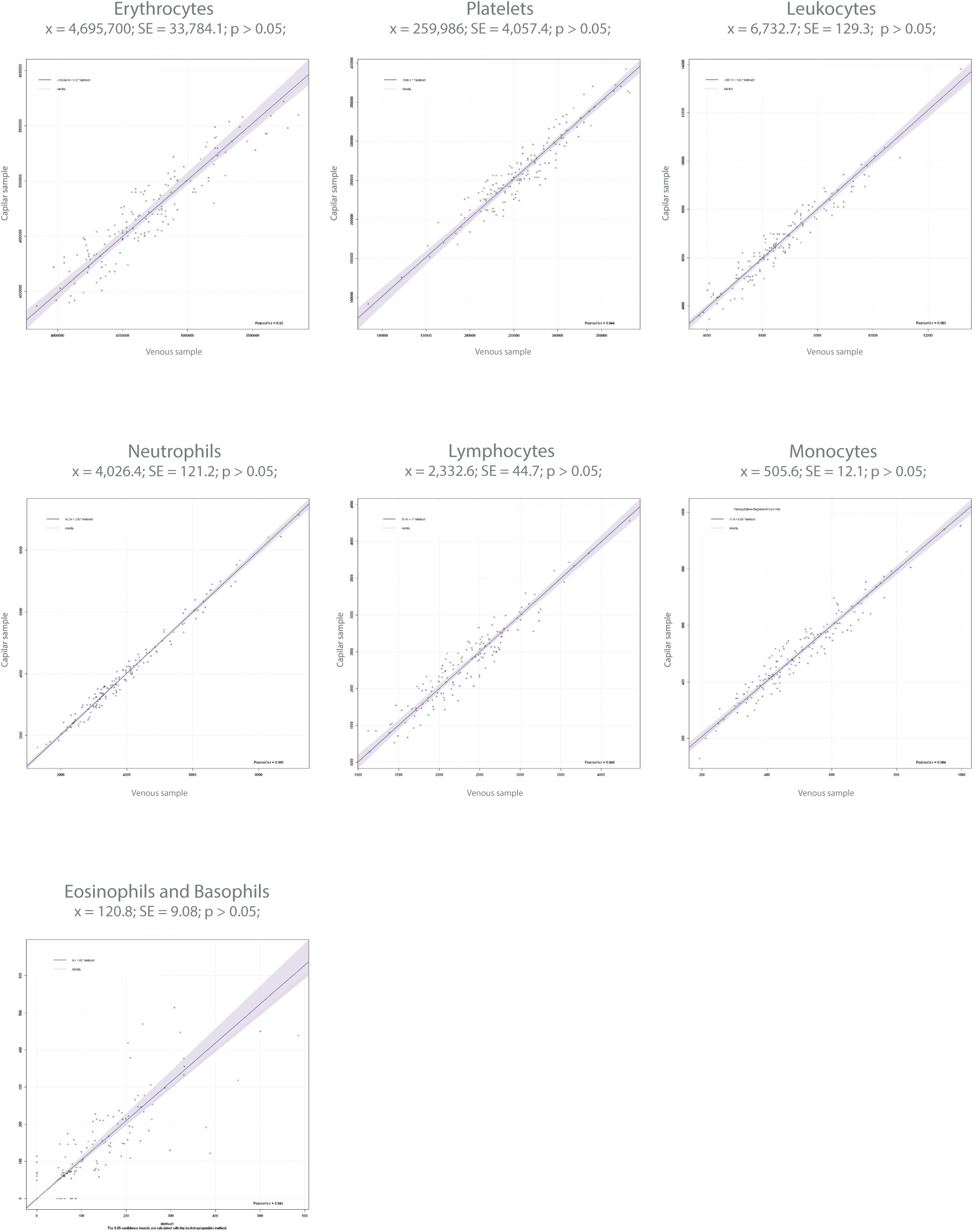
Anticoagulant influence and the effect of sample type. Graphs indicate the Hilab system results for venous (plus K_3_EDTA) x fingerprint blood samples. Mean 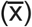, standard deviation (SD), and p-values of Paired Student T-test are demonstrated for each analyte.

### 4. Flagging Study

For RBC morphological abnormalities, 54 positive and 396 negative samples were analyzed. These analyses covered cases of microcytosis, anisocytosis, and macrocytosis. As a result, high accuracy (97.06%), sensitivity (97.06%), specificity (100.0%), and balanced accuracy (98.1%) were observed. For PLT clumps, 17 positive and 433 negative samples were analyzed. Even with a minor number of positive samples, perfect accuracy (100.0%), sensitivity (100.0%), specificity (100.0%), and balanced accuracy were observed (100.0%). Finally, evaluating the presence of immature cells from both red and white lineage, 10 positive and 440 negative samples were analyzed. As shown above, high accuracy (95.73%), sensitivity (95.73%), specificity (100%), and balanced accuracy (89.9%) were observed (Table 3). Also, all evaluated parameters demonstrated strong kappa coefficients (RBC morphology - 0.98; PLT - 1.00; Immature cells - 0.89).

**Table 3.** Accuracy, specificity, sensibility, kappa coefficient, and balanced accuracy of the flagging study, comparing the Hilab system to the microscopy technique.

| **Disturbance** | **Accuracy (%)** | **Specificity (%)** | **Sensitivity (%)** | **Kappa** | **Balanced Accuracy (%)** |
| --- | --- | --- | --- | --- | --- |
| **RBC morphology** | 97.06 | 100.0 | 97.06 | 0.98 | 98.1 |
| **PLT** | 100.0 | 100.0 | 100.0 | 1.00 | 100.0 |
| **Immature cells** | 95.73 | 100.0 | 95.73 | 0.89 | 89.9 |

## Discussion

Most handheld CBC devices commercially available present high costs and are not liable to calibration or control procedures, which results in poor quality compared to standard instruments ^11,12^. Our study provided an extensive clinical validation of the Hilab system to CBC point-of-care test, evaluating parameters like comparability, precision, and flagging studies. Over the entire measuring range, all values provided by this new approach presented high sensibility, specificity, and accuracy, compared to a sophisticated hematological analyzer (Sysmex XE-2100). Also, a high correlation was observed for all parameters evaluated. Thus, considering the need for blood count point-of-care tests, especially for quickly patient management, the study indicated that the Hilab system provides fast, accurate, low cost, and robust blood cell analysis for reliable clinical use.

The comparability study encompassed a significant range of values to all analytes, evaluating different health conditions. The robust values provided by the Hilab system, compared to the conventional hematological analyzer, can be observed by high Pearson correlation values (≥ 0.8), low bias, and the absence of statistically significant differences (p > 0.05) to most analytes (Figure 2). Also, strong values (≥ 0.8) of accuracy, specificity, sensitivity, and balanced accuracy were obtained. The kappa coefficient results emphasize the reliable results of the Hilab System (≥ 0.8; Table 1), demonstrating a strong level of data agreement between these two different CBC methodologies.

The EOS/BAS count was the only parameter that demonstrated differences statistically significant (p < 0.05) between Sysmex XE-2100 and Hilab results. Considering that the EOS and BAS are the less prevalent WBC subpopulation, the lower evaluation area (1 mm^2^) of the Hilab Lens device compared to Sysmex XE-2100 may influence these cell counts. In this sense, previous studies ^13^ demonstrate that even considering conventional hematological analyzers, the slightest area evaluations differences result in differences in EOS and BAS quantifications. However, it’s important to emphasize that considering the clinical range of these analytes, great values of accuracy, sensibility, specificity, and balanced accuracy were acquired (≥ 0.8; Table 1) with the Hilab system, as well as the kappa coefficient (> 0.8).

A special note should be taken regarding WBCs differentiation into four subpopulations (NEU, LINF, MONO, and EOS/BAS). Although other CBC devices provide 5-part WBC differentiation, the Hilab system can supply detailed patient health information, considering that 3-part hematology analyzers already provide enough information for most clinical settings. Also, in case of EOS/BAS count increase, the clinical report of patients can easily distinguish which cell subpopulation is changed.

The precision assay showed that all RBC, WBC, PLT, and HB levels presented the CV and SD values within limits proposed by ELFM (Table 2). These data demonstrate the high repeatability and reproducibility of the Hilab system. Regarding others CBC analytes, in this assay, it was chosen not to evaluate the 4-part differential WBC analytes, based on the incompatibility between commercial hematological controls and the Hilab reagents. Also, as HT, MCV, and MCH are estimated by HB and RBC values, these parameters were not regarded in this analysis.

As previously observed ^5^, comparing the results of blood samples collected from venous (plus K_3_EDTA anticoagulant) or by finger stick, none cell analytes presented statistically significant differences (p > 0.05; paired T-test; Figure 3) among the collection methods. Therefore, the blood collection method of the Hilab system was validated. Furthermore, this data demonstrated that the anticoagulant K_3_EDTA does not interfere with the Hilab system result. Although hematimetric parameters have not been evaluated in this analysis, other authors have already shown that no statistically significant differences are found between venous and capillary samples for these analytes ^14^. Also, tests demonstrated the absence of K_3_EDTA interference in HB results (data not shown).

Finally, based on the fact that the Hilab System uses the microscopy technique, which is considered the gold standard method for cell identification, the flagging study focused on the analysis of main CBC test alterations: RBC morphological variation, PLT clumps, and the presence of immature cells. For all alterations evaluated (Table 3), a high correlation between the Hilab system and the manual microscopy technique associated with trained hematologists was demonstrated. Considering the elevated costs associated with trained laboratory technicians, associated with the high time spent in individual blood smears analysis, the fast and precise flagging analysis of the Hilab device must be considered a relevant advantage of this hematological POCT.

## Data Availability

For original data and analysis code, please contact.

## Acknowledgments

This study was supported by research funding from Hi Technologies, to M. F. and S. R.

## Authorship Contributions

All authors contributed equally to the manuscript, realizing a) significant contribution to design, data interpretation, acquisition, and/or analysis; b) Investigation of the intellectual content of the article; and c) writing of the original manuscript. Besides, the M. F., S. R., I. S., F. T., and R.S fabricated the Hilab System. V. R, E. S, M. C., and F. S. made the deep learning approach for both cell detection and classification. The blood sample testing and data analysis was performed by A. G., C. A., M. P., T. J., and E. S. The J. S. and B. A. oversaw the entire project.

